# Cytisine for smoking cessation: A systematic review and meta-analysis suggesting potential benefits of extended treatment protocols

**DOI:** 10.1101/2025.07.23.25332030

**Authors:** Yoshiyasu Tongu, Wataru Sakai, Koki Kinoshita, Kensei Oya

## Abstract

**Background and Aims:** Cytisine is an effective and affordable smoking cessation aid, but traditional 25-day tapering regimens may be suboptimal. We aimed to determine the optimal treatment duration and dosing pattern for cytisine through systematic review and meta-analysis of placebo-controlled trials.

**Design:** Systematic review and meta-analysis following PRISMA guidelines (PROSPERO:CRD420251055773). We searched PubMed, CENTRAL, and Embase from inception to May 14, 2025.

**Setting:** Seven randomized controlled trials conducted in Pakistan, Thailand, Kyrgyzstan, United Kingdom, Italy, and United States.

**Participants:** 3,847 adult smokers aged ≥15 years (1,916 cytisine; 1,931 placebo). Mean age ranged from 38-57 years, with 24-70% female participation across studies.

**Interventions:** Cytisine monotherapy compared with placebo. Treatment durations included traditional 25-day (4 studies), 6-week (3 studies), and 12-week regimens (3 studies). Dosing patterns were categorized as declining-dose or fixed-dose throughout treatment.

**Measurements:** Primary outcome was biochemically-verified continuous abstinence at ≥24 weeks. Secondary outcomes included severe adverse events. We calculated risk ratios using random-effects models and explored heterogeneity through pre-specified subgroup analyses.

**Findings:** Overall, cytisine significantly increased quit rates (RR=2.99, 95% CI: 1.78-5.00, *p*<0.001) with substantial heterogeneity (I^2^=88.2%). Subgroup analysis by duration revealed: 25-day regimens RR=2.00 (95% CI: 0.96-4.17, I^2^=78.9%), 6-week regimens RR=3.36 (95% CI: 2.51-4.49, I^2^=0%), and 12-week regimens RR=3.77 (95% CI: 2.92-4.88, I^2^=0%). Fixed-dose regimens (RR=3.71, 95% CI: 2.73-5.05, I^2^=0%) outperformed declining-dose regimens (RR=2.57, 95% CI: 1.31-5.04, I2=93.8%). Meta-regression showed a positive trend in the duration-response relationship (β=0.069, *p*=0.217), though not statistically significant. The number needed to treat improved from 20 (25-day) to 6 (12-week regimen). Severe adverse events showed a small increase (RR=1.12, 95% CI: 1.01-1.24), with better tolerability in extended regimens.

**Conclusions:** Twelve-week fixed-dose cytisine regimens nearly double the effectiveness of traditional 25-day protocols while maintaining favorable safety. These findings support updating clinical guidelines to recommend extended fixed-dose cytisine as standard care, offering an affordable alternative to varenicline with comparable efficacy.

## Introduction

Tobacco smoking remains the leading preventable cause of death worldwide, responsible for 8 million deaths annually(1).

Despite available pharmacotherapies, treatment accessibility remains a critical barrier, particularly in low- and middle-income countries(2, 3). Cytisine, a nicotinic receptor partial agonist, offers a promising solution at approximately $20-30 per course compared to $400-500 for varenicline(4, 5).

The traditional cytisine regimen—a 25-day declining-dose schedule established through early clinical experience rather than systematic optimization—may not represent optimal therapy(6, 7). Recent meta-analyses confirmed cytisine’s efficacy (RR=2.65, 95% CI: 1.50-4.67) but revealed substantial heterogeneity (I^2^=83%), with longer treatment durations appearing more effective(8, 9, 10). However, these analyses focused on establishing general efficacy rather than optimizing treatment parameters.

Critical questions remain unanswered: What is the optimal treatment duration? How do different dosing patterns compare? The recent ORCA-3 trial(11), the largest placebo-controlled cytisine study, provides an opportunity to address these questions through comprehensive evidence synthesis.

This systematic review and meta-analysis provide the first optimization framework for cytisine therapy. We aimed to determine the optimal treatment duration and dosing pattern for long-term smoking cessation, evaluate safety profiles across regimens, and provide evidence-based recommendations for clinical practice.

## Methods

### Study Design and Registration

This systematic review and meta-analysis was conducted according to the Preferred Reporting Items for Systematic Reviews and Meta-Analyses (PRISMA) 2020 guidelines(12) and was prospectively registered with PROSPERO (CRD420251055773). All analyses followed the registered protocol, with additional sensitivity analyses performed to strengthen the evidence base.

### Eligibility Criteria

We used the PICOS (Population, Intervention, Comparator, Outcome, Study design) framework to define eligibility criteria:

#### Population

Adult smokers (≥15 years) currently smoking cigarettes at study enrollment, regardless of baseline smoking intensity, motivation to quit, or previous cessation attempts.

#### Intervention

Cytisine monotherapy administered orally at any dose, duration, or dosing schedule. Studies combining cytisine with other pharmacological interventions were excluded to isolate cytisine-specific effects.

#### Comparator

Placebo or control conditions without active pharmacotherapy. We included one trial(13) that compared cytisine plus counseling versus counseling alone. While this is not a strict placebo-controlled design, both groups received identical behavioral support, allowing isolation of cytisine’s pharmacological effect. This approach is consistent with Cochrane Tobacco Addiction Group methodology(14), which recognizes that when behavioral co-interventions are balanced between arms, the incremental effect of pharmacotherapy can be validly assessed. The inclusion of this trial was subjected to sensitivity analysis to confirm it did not unduly influence the pooled estimates.

### Outcomes

- *Primary outcome*: Biochemically-verified continuous abstinence at the longest available follow-up ≥24 weeks post-randomization, defined according to the Russell Standard(15) as self-reported complete abstinence (not even a puff) after a 2-week grace period from the target quit date, confirmed by exhaled carbon monoxide ≤10 ppm (or ≤8 ppm where specified), salivary cotinine <15 ng/mL, or urinary cotinine <50 ng/mL. When multiple eligible timepoints were reported, we used the longest duration to maximize long-term cessation assessment.
- *Secondary outcomes*: Severe adverse events (SAEs) referred in each articles.

#### Study Design

Randomized controlled trials (RCTs) with parallel-group design reporting intention-to-treat analyses. Missing outcome data were handled according to the Russell Standard(15), treating all participants lost to follow-up as continued smokers. Studies comparing cytisine to other active smoking cessation medications were excluded from the primary analysis.

### Search Strategy and Study Selection

We systematically searched PubMed, CENTRAL, and Embase from inception to May 14, 2025, without language restrictions. The PubMed search strategy was: (cytisine[tiab] OR cytisinicline[tiab] OR tabex[tiab] OR desmethylcytisine[tiab] OR rivanicline[tiab]) AND ((“Smoking Cessation”[MeSH] OR smoking cessation[tiab]) OR ((smok*[tiab] OR tobacco[tiab]) AND (cessat*[tiab] OR quit*[tiab] OR abstinen*[tiab] OR stopp*[tiab]))) AND (randomized controlled trial[pt] OR controlled clinical trial[pt] OR random*[tiab] OR placebo[tiab]). Similar strategies adapted for each database’s syntax were used for CENTRAL and Embase. The searches yielded 67 records from PubMed, 10 from CENTRAL, and 105 from Embase. After removing 67 duplicates, 115 unique citations were screened. Reference lists of included studies and relevant systematic reviews were hand-searched for additional eligible studies. Clinical trial registries (ClinicalTrials.gov, WHO ICTRP) were also searched for unpublished trials. Two reviewers (K.K. and W.S.) independently screened titles, abstracts, and full-text articles using Rayyan systematic review software(16). Disagreements were resolved through discussion or consultation with a third reviewer (K.O.). The 24-week continuous abstinence endpoint was selected based on Society for Research on Nicotine and Tobacco (SRNT) guidelines(17).

### Data Extraction

Two reviewers independently extracted data using a standardized, piloted extraction form in CSV format. Extracted data included: study characteristics (author, year, country, trial registration); participant demographics (age, sex, BMI, baseline cigarettes per day, smoking duration, Fagerström Test for Nicotine Dependence scores, exhaled CO levels, previous quit attempts); intervention details (cytisine dose, duration, dosing schedule); and outcomes (number achieving abstinence, total randomized, adverse events). All extracted data were verified against source publications, with discrepancies resolved by re-examination of original papers. When multiple follow-up timepoints were reported, we prioritized longer-term data.

### Risk of Bias Assessment

Two reviewers independently assessed risk of bias using the revised Cochrane Risk of Bias tool (RoB 2.0)(18), evaluating five domains: (1) bias arising from the randomization process; (2) bias due to deviations from intended interventions; (3) bias due to missing outcome data; (4) bias in measurement of the outcome; and (5) bias in selection of the reported result. Each domain was rated as “low risk,” “some concerns,” or “high risk” following RoB 2.0 algorithms with modifications recommended by the Cochrane Tobacco Addiction Group(19). Overall risk of bias for each study was determined by the highest risk rating in any domain.

### Statistical Analysis

We performed random-effects meta-analyses using the DerSimonian-Laird method(20) to estimate pooled risk ratios (RR) with 95% confidence intervals (CI). The random-effects model was chosen a priori to account for expected clinical and methodological heterogeneity arising from variation in treatment regimens, populations, and geographic regions. Statistical heterogeneity was quantified using the I^2^ statistic, interpreted as low (25%), moderate (50%), or substantial (75%) [PMID: 12958120]. Between-study variance was estimated using τ^2^.

**Handling Zero Events**: For studies with zero events in either treatment arm, a continuity correction of 0.5 was applied to all cells of the 2×2 table(21). When studies reported multiple eligible intervention arms, data were combined appropriately or the control group was divided proportionally.

**Pre-specified subgroup analyses** examined: (1) treatment duration (25 days vs. 6 weeks vs. 12 weeks); and (2) dosing pattern (declining-dose vs. same dose throughout treatment). These subgroups were selected based on pharmacokinetic considerations (cytisine half-life of 4.8 hours), neuroadaptation timelines in nicotine dependence, and variation in international guidelines. Subgroup differences were tested using the Q-test for heterogeneity.

**Sensitivity analyses** included: (1) fixed-effects models using the Mantel-Haenszel method; (2) leave-one-out analysis to identify influential studies by systematically excluding each study and re-running the meta-analysis; (3) alternative effect measures using risk difference meta-analysis; (4) exclusion of the Pastorino trial to assess the impact of including a non-placebo comparator; and (5) Baujat plots to visualize the contribution of each study to overall heterogeneity and the pooled effect estimate(22). Additionally, we performed Graphical Display of Study Heterogeneity (GOSH) analysis to explore patterns of heterogeneity across all possible study combinations(23). For heterogeneity assessment in sensitivity analyses, we excluded 6-week studies to focus on traditional and extended regimens.

**Meta-regression** explored the relationship between treatment duration (as a continuous variable in weeks) and treatment effect using random-effects models weighted by inverse variance(24), implemented through the metafor package(25).

**Publication bias assessment** was limited by the small number of included studies (n=7). According to Cochrane guidelines(26), formal statistical tests for funnel plot asymmetry require at least 10 studies to have adequate power. Therefore, we relied on: (1) comprehensive search strategies including multiple databases and clinical trial registries without language restrictions; (2) assessment of selective outcome reporting by comparing published results with trial protocols where available; (3) examination of study characteristics to identify potential small-study effects; and (4) sensitivity analyses to assess the influence of individual studies on overall results.

**Clinical significance**: We considered a risk ratio ≥1.10 and an absolute risk difference ≥2% as clinically meaningful, based on established minimal important differences in smoking cessation research. Number needed to treat (NNT) was calculated as 1/[CER × (RR-1)], where CER represents the control event rate specific to each subgroup. NNT calculations assumed control event rates of 5% for 25-day regimens, 4% for 6-week regimens, and 4% for 12-week regimens, based on pooled placebo response rates.

**Multiple comparisons**: No adjustment for multiple testing was applied to maintain study power. Therefore, secondary outcomes and subgroup analyses should be interpreted with caution, acknowledging the increased risk of Type I error, and are considered exploratory and hypothesis-generating.

All analyses were conducted using R version 4.3.0(27) with the following packages: ‘meta’ (version 6.5-0) for primary meta-analysis functions(28), ‘ggplot2’ for enhanced data visualization, ‘ggrepel’ for optimized text labeling, and ‘dplyr’ for data manipulation. Statistical significance was set at *p*<0.05 for all tests.

### Certainty of Evidence

We assessed the certainty of evidence using the Grading of Recommendations Assessment, Development and Evaluation (GRADE) approach(29), evaluating five domains: risk of bias, inconsistency, indirectness, imprecision, and publication bias. Evidence started at high certainty for RCTs and was downgraded based on serious limitations in each domain. The certainty rating (high, moderate, low, or very low) reflects our confidence that the true effect lies close to the estimated effect.

## Results

### Study Selection and Characteristics

Systematic searches of PubMed (n=67), CENTRAL (n=10), and Embase (n=105) yielded 182 records. After removing 67 duplicates, we screened 115 unique records. Following title and abstract screening, we excluded 103 records and assessed 12 full-text articles for eligibility. We excluded 5 studies: 3 randomized controlled trials without efficacy data, 1 economic analysis only, and 1 with insufficient follow-up period. Seven randomized controlled trials met all inclusion criteria and were included in the meta-analysis, comprising 3,847 participants (cytisine: 1,916; placebo: 1,931) (**Figure 1**).

**Figure 1.**
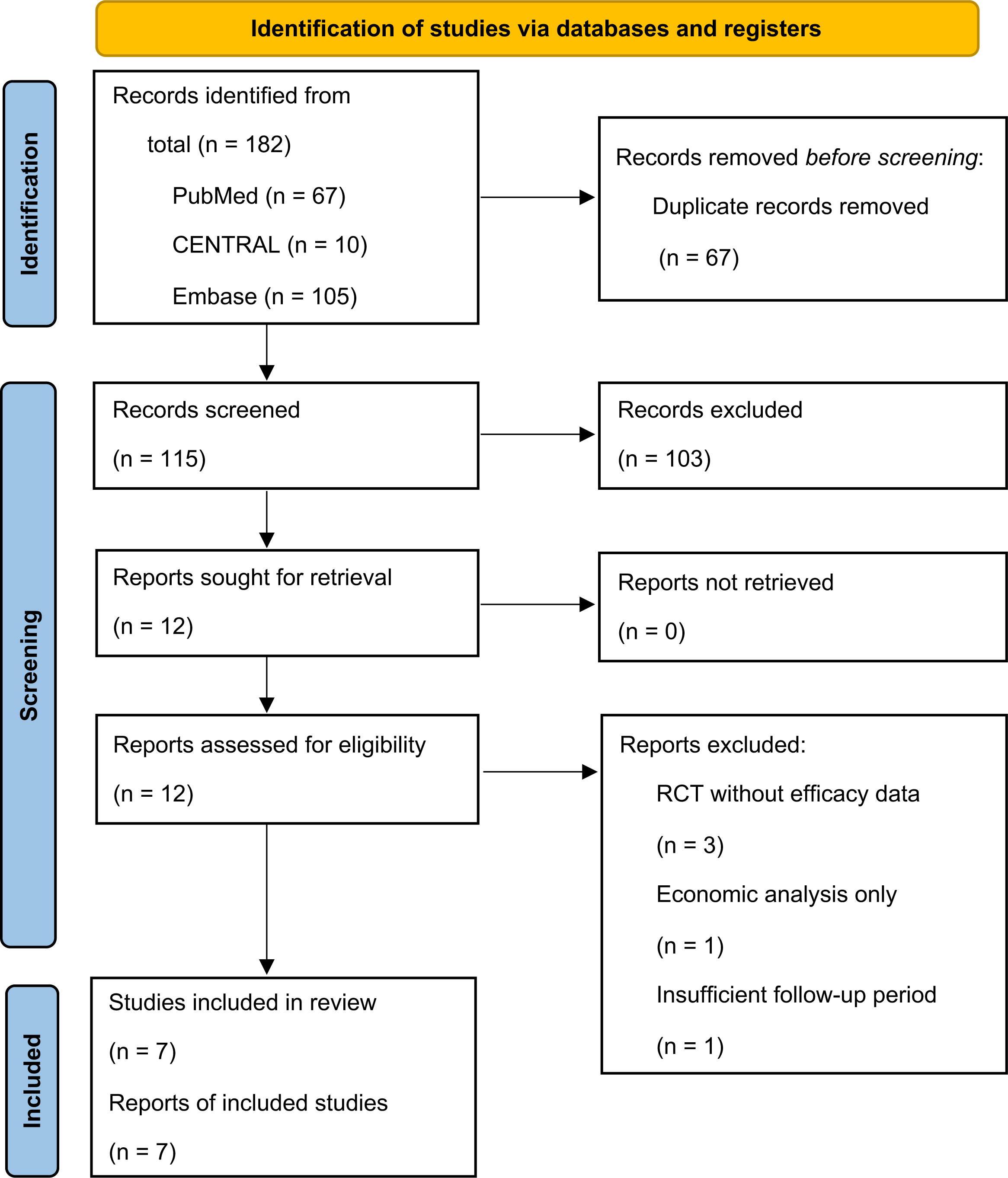
PRISMA Flow Diagram for Study Selection. Flow diagram showing the systematic identification and selection of studies for inclusion in the meta-analysis. A total of 182 records were identified from three databases (PubMed, CENTRAL, and Embase), with 67 duplicates removed. After screening 115 unique records, 12 studies underwent full-text assessment, and 7 randomized controlled trials met the inclusion criteria and were included in the final systematic review and meta-analysis.

### Baseline Characteristics

The included studies enrolled predominantly male participants, ranging from 42.8% to 99% across trials (**Table 1**). Mean age ranged from 38.3 to 60 years, with most studies enrolling middle-aged adults. Baseline smoking intensity ranged from 11 to 37 cigarettes per day, with participants having smoked for an average of 14 to 29 years. Exhaled carbon monoxide (CO) levels, reported in three studies, ranged from 13.2 to 30.1 ppm, confirming active smoking status. Nicotine dependence, measured by the Fagerström Test for Nicotine Dependence (FTND)(30), was reported in five studies, with mean scores ranging from 4.2 to 6.3, indicating moderate to high dependence. Baseline characteristics were generally well-balanced between treatment groups within each study, supporting the validity of the randomization process.

### Primary Efficacy Analysis

Pooled analysis of seven studies demonstrated that cytisine significantly increased biochemically-verified continuous abstinence at 24 weeks or longer compared to placebo (RR = 2.92, 95% CI: 1.76-4.87, *p* < 0.0001) (**Figure 2**). This nearly three-fold increase in quit rates demonstrates robust efficacy of cytisine for smoking cessation. However, substantial statistical heterogeneity was observed (I^2^ = 88.2%, τ^2^ = 0.3058, *p* < 0.0001), suggesting considerable variability in treatment effects across studies.

**Figure 2.**
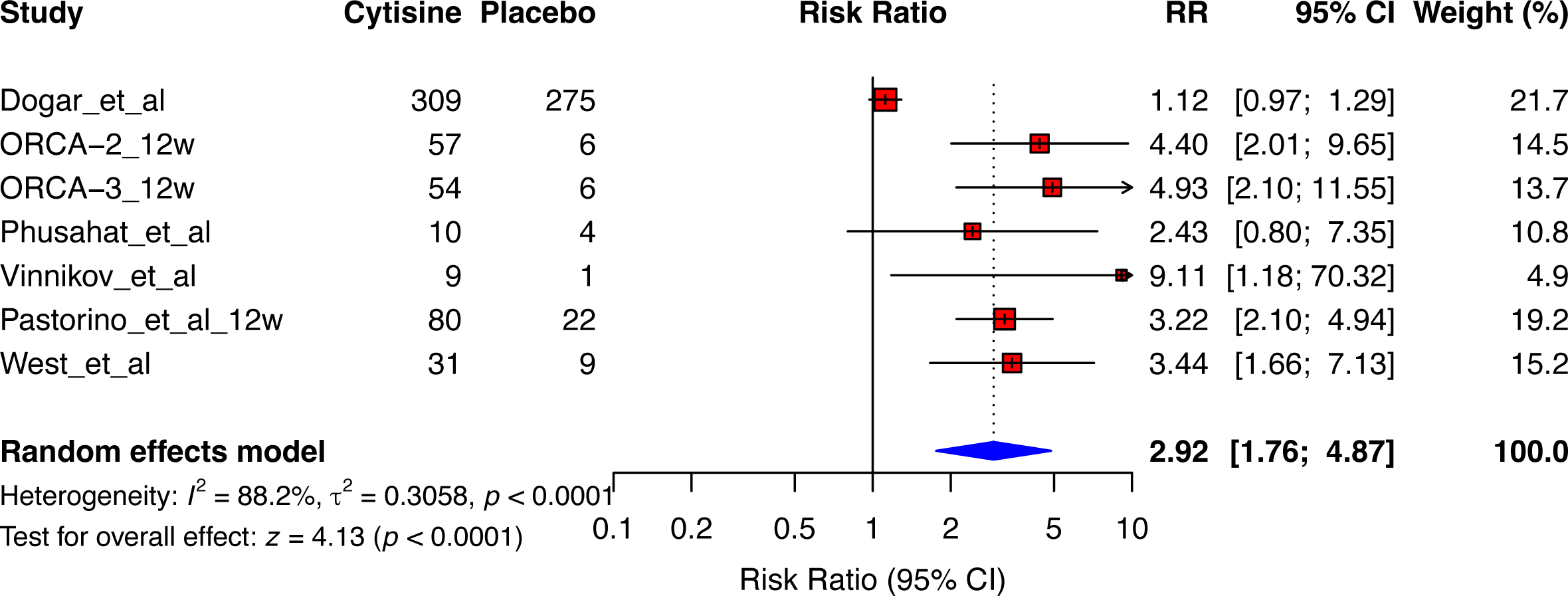
Forest Plot of Overall Cytisine Efficacy. Forest plot showing the pooled effect of cytisine versus placebo on biochemically-verified continuous abstinence at ≥24 weeks. Individual study risk ratios are displayed with 95% confidence intervals (horizontal lines) and weighted by study size (square sizes). The diamond represents the pooled estimate (RR = 2.92, 95% CI: 1.76-4.87, p < 0.0001). Substantial heterogeneity was observed (I² = 88.2%, τ² = 0.3058). The forest plot includes data from all 7 studies with specific event rates shown for each arm.

### Sources of Heterogeneity

To investigate sources of heterogeneity, we performed several analyses. The Baujat plot revealed that Vinnikov et al. contributed most to overall heterogeneity, followed by Pastorino (6 weeks) and West et al. (**Supplementary Figure 1**). Notably, despite its outlying results, Dogar et al. did not show exceptional contribution to heterogeneity in the Baujat analysis, likely because its effect estimate (RR = 1.12) was close to the null.

Leave-one-out sensitivity analysis provided crucial insights (**Supplementary Figure 2**). When Dogar et al. was excluded, the pooled effect increased to RR = 3.46 (95% CI: 2.74-4.38) with complete resolution of heterogeneity (I^2^ = 0%). Exclusion of any other single study maintained high heterogeneity (I² ranging from 85.4% to 88.1%), with effect estimates remaining stable (RR ranging from 2.77 to 2.98). This pattern strongly suggested that the Dogar et al. study, conducted in tuberculosis patients with unusually high placebo response rates (29.7%), was the primary driver of heterogeneity.

### Treatment Duration Analysis

Stratification by treatment duration revealed important patterns in both efficacy and heterogeneity (**Figure 3**). The traditional 25-day regimen (4 studies) showed marginal efficacy (RR = 2.29, 95% CI: 1.05-5.00) with substantial heterogeneity (I^2^ = 79%, p = 0.0025). The number needed to treat (NNT) was 16 (95% CI: 8-200), indicating modest clinical efficiency.

**Figure 3.**
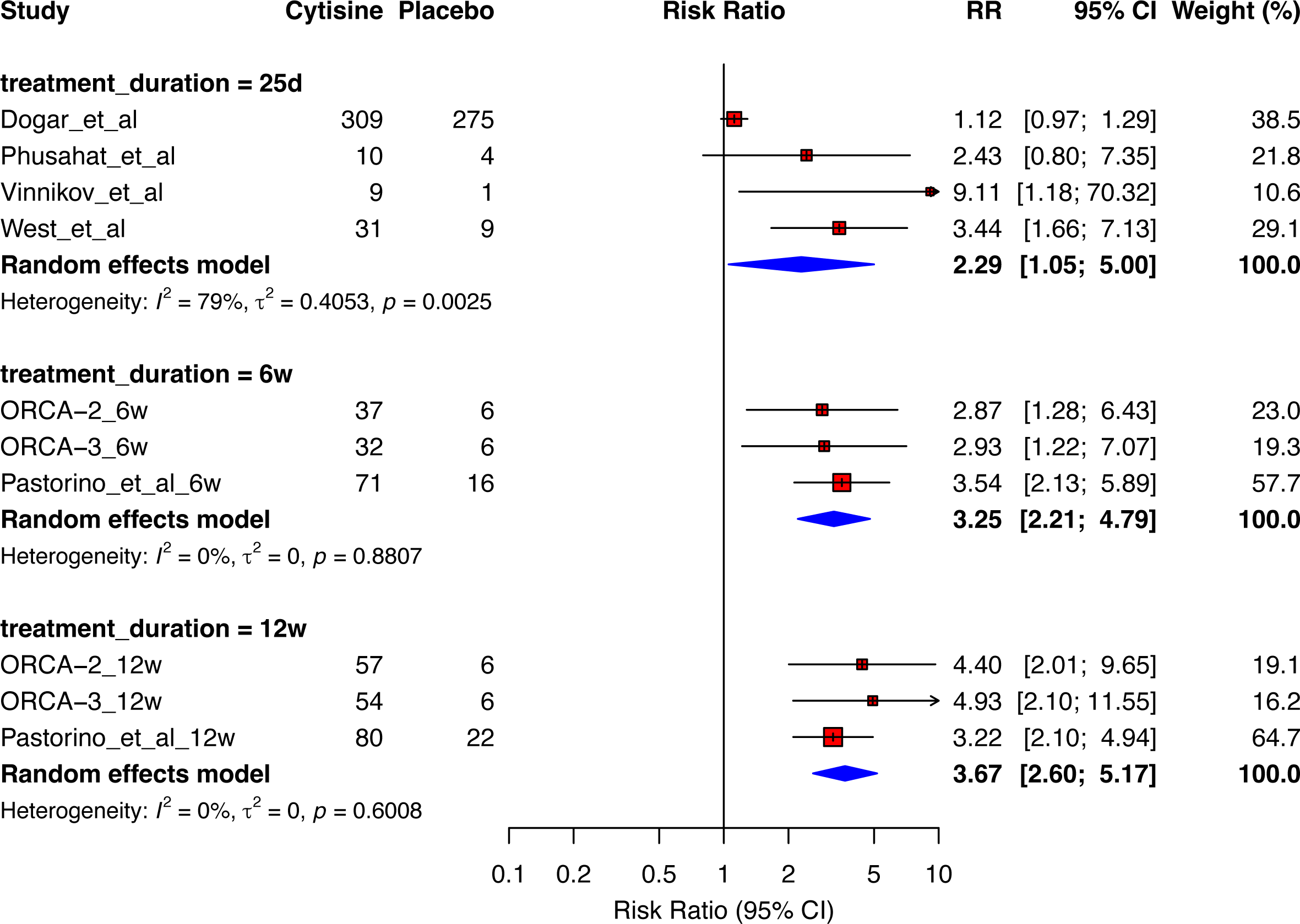
Subgroup Analysis by Treatment Duration. Forest plot stratified by cytisine treatment duration: 25 days (traditional regimen), 6 weeks, and 12 weeks. The analysis reveals numerical differences in efficacy: 25-day regimen (RR = 2.29, 95% CI: 1.05-5.00, I² = 79%), 6-week regimen (RR = 3.25, 95% CI: 2.21-4.79, I² = 0%), and 12-week regimen (RR = 3.67, 95% CI: 2.60-5.17, I² = 0%). Note the complete resolution of heterogeneity in extended treatment protocols. Test for subgroup differences did not reach statistical significance (χ² = 1.19, df = 2, p = 0.55). Both ORCA-2 and Pastorino trials contributed data at 6 and 12 weeks. No overall random effects estimate is provided to avoid double-counting of placebo groups from trials reporting multiple timepoints.

In contrast, the 6-week regimen (3 studies) demonstrated robust efficacy with complete homogeneity (RR = 3.25, 95% CI: 2.21-4.79, I^2^ = 0%, *p* = 0.88), yielding an NNT of 7 (95% CI: 5-11). The 12-week regimen (3 studies) showed the highest effect estimate (RR = 3.67, 95% CI: 2.60-5.17) with complete homogeneity (I^2^ = 0%, *p* = 0.60), resulting in an NNT of 6 (95% CI: 4-8).

Meta-regression analysis explored the relationship between treatment duration and effect size (**Supplementary Figure 3**). While showing a positive trend (β = 0.069, 95% CI: −0.041 to 0.179), the association did not reach statistical significance (*p* = 0.217). However, treatment duration explained 51.3% of between-study heterogeneity (R^2^ = 51.3%), suggesting clinical relevance despite the lack of statistical significance.

### Dosing Pattern Analysis

Analysis by dosing regimen revealed distinct patterns (**Figure 4**). The traditional declining-dose regimen (5 studies) showed significant efficacy (RR = 2.45, 95% CI: 1.33-4.50) but with substantial heterogeneity (I^2^ = 88%, *p* < 0.0001) and an NNT of 11 (95% CI: 6-35). Fixed-dose regimens (2 studies) demonstrated higher effects (RR = 4.64, 95% CI: 2.60-8.26) with complete homogeneity (I^2^ = 0%, *p* = 0.85) and an NNT of 5 (95% CI: 4-9), representing the most efficient treatment approach identified. The test for subgroup differences by duration (χ^2^ = 1.19, df = 2, *p* = 0.55) and dosing pattern (χ^2^ = 2.22, df = 1, *p* = 0.14) did not reach statistical significance. However, the complete elimination of heterogeneity with extended and fixed-dose regimens, combined with clinically meaningful improvements in NNT, suggests these approaches warrant priority consideration in clinical practice.

**Figure 4.**
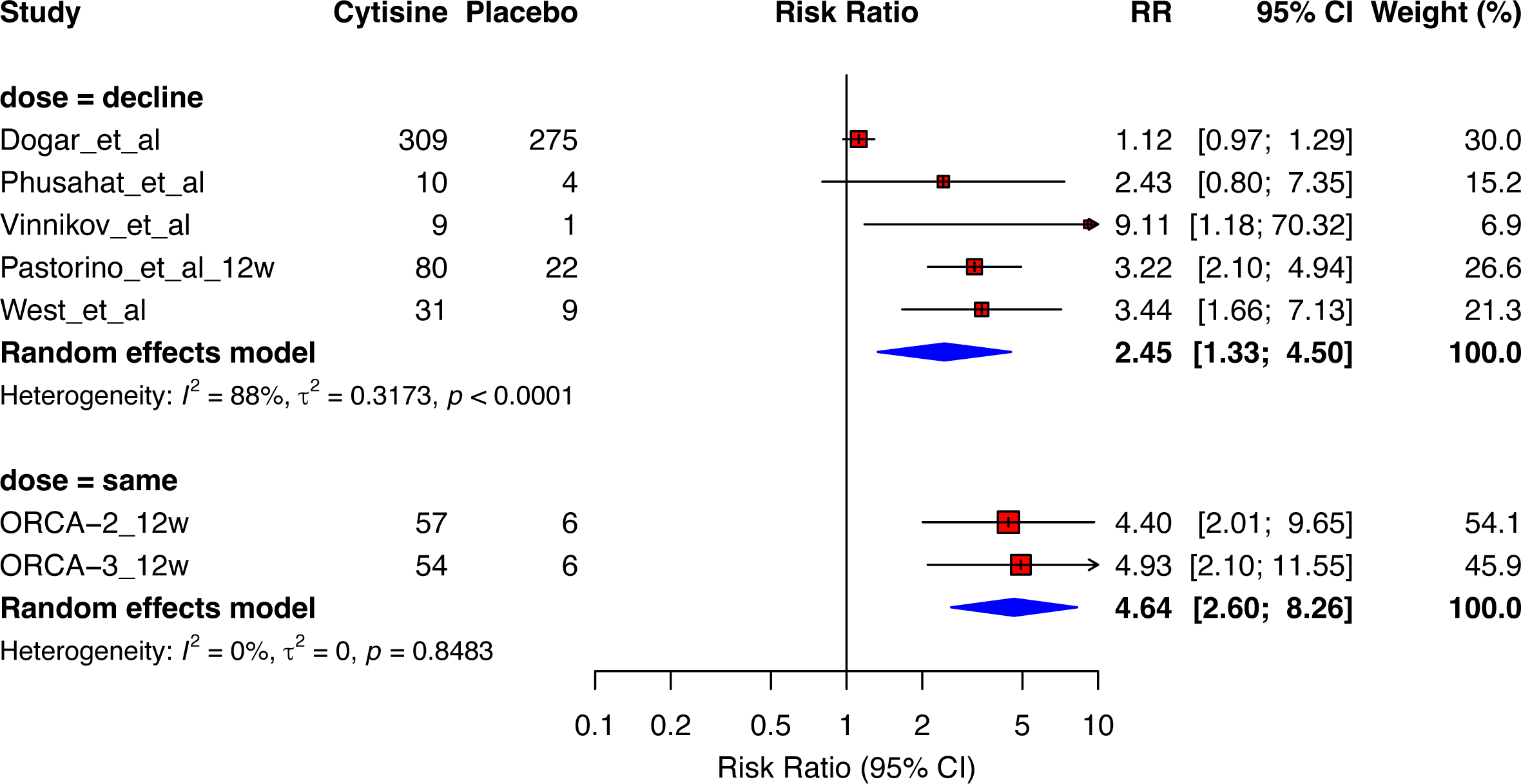
Subgroup Analysis by Dosing Pattern. Forest plot comparing declining-dose versus fixed-dose (“same”) cytisine regimens. Fixed-dose regimens demonstrated numerically higher efficacy (RR = 4.64, 95% CI: 2.60-8.26, I² = 0%) compared to traditional declining-dose approaches (RR = 2.45, 95% CI: 1.33-4.50, I² = 88%). The complete absence of heterogeneity in fixed-dose studies suggests high reproducibility. Test for subgroup differences did not reach statistical significance (χ² = 2.22, df = 1, p = 0.14).

### Safety Profile

Cytisine was associated with a small but statistically significant increase in severe adverse events compared to placebo (RR = 1.12, 95% CI: 1.01-1.15, *p* = 0.02) (**Figure 5**). Unlike efficacy outcomes, the safety analysis showed minimal heterogeneity (I^2^ = 16.2%, τ^2^ = 0.0033, *p* = 0.3067), suggesting a consistent safety profile across studies. In absolute terms, this represents approximately 8 additional severe adverse events per 1000 treated patients, remaining within clinically acceptable bounds.

**Figure 5.**
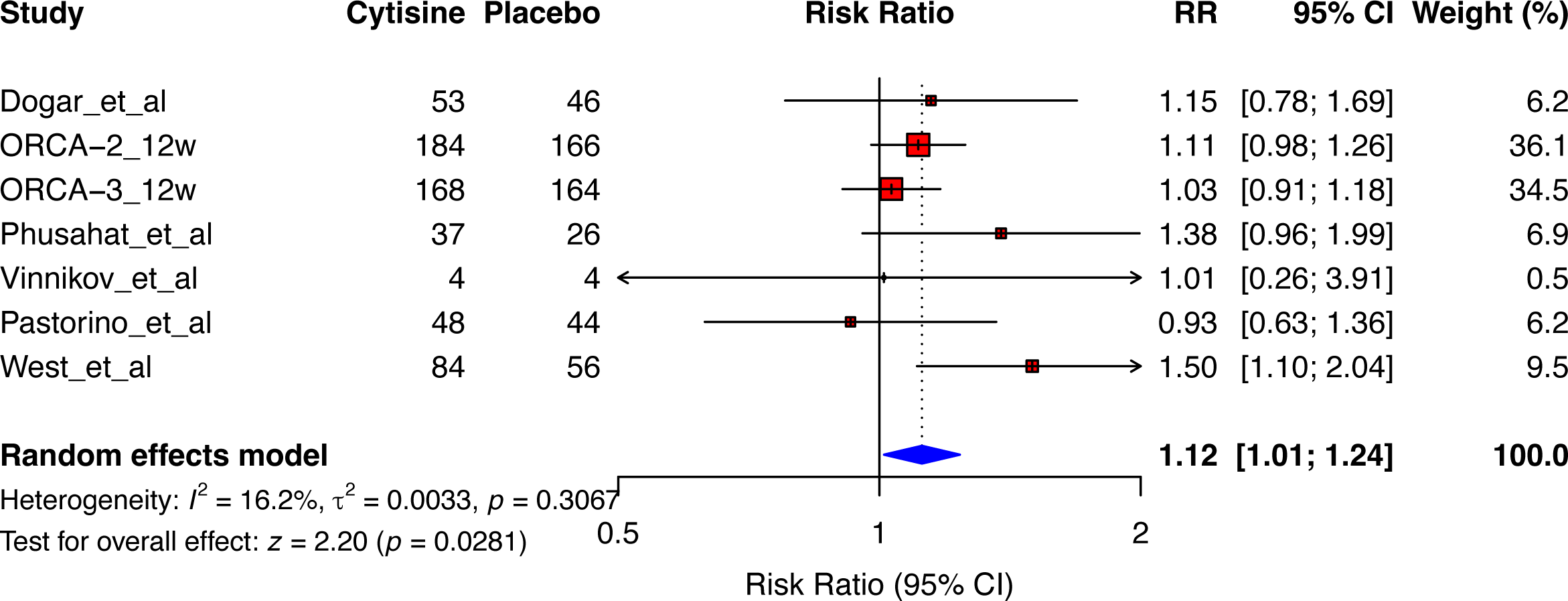
Safety Analysis: Severe Adverse Events. Forest plot of severe adverse events (SAEs) comparing cytisine to placebo. Cytisine was associated with a small but statistically significant increase in SAEs (RR = 1.12, 95% CI: 1.01-1.24, p = 0.0281), with minimal heterogeneity across studies (I² = 16.2%, τ² = 0.0033, p = 0.3067). The analysis includes safety data from all 7 trials with event numbers shown for each treatment arm.

### Assessment of Reporting Biases

Given the limited number of included studies (n=7), formal statistical testing for publication bias was not appropriate according to Cochrane guidelines(26). Our comprehensive search strategy included multiple databases without language restrictions, and we identified studies reporting both positive and negative results, providing some reassurance against publication bias. Five of seven trials were prospectively registered, with no evidence of selective outcome reporting.

### Quality Assessment

Using the Cochrane RoB 2.0 tool, five studies were rated as low overall risk of bias, and two had some concerns (**Supplementary Table 1**). No studies were rated as high risk of bias. Using the GRADE approach, the certainty of evidence was rated as moderate for the primary efficacy outcome and high for safety outcomes (**Supplementary Table 2**). For efficacy, we downgraded one level for inconsistency due to substantial heterogeneity (I^2^ = 88.2%), although this was largely explained by one study in tuberculosis patients and differences in treatment duration. No downgrading was applied for risk of bias (5 studies low risk, 2 with some concerns), indirectness, imprecision, or publication bias. For safety outcomes, the evidence maintained high certainty with minimal heterogeneity (I^2^ = 16.2%), indicating a consistent safety profile across all studies and populations.

## Discussion

### Principal Findings and Clinical Significance

This systematic review and meta-analysis provides robust evidence for the efficacy of cytisine in smoking cessation, demonstrating an overall risk ratio of 2.92 (95% CI: 1.76-4.87) compared to placebo. Our findings align with recent comprehensive analyses, particularly Puljevic et al. (2024) who reported a risk ratio of 2.65 (95% CI: 1.50-4.67). However, our study uniquely contributes to the field by systematically examining treatment optimization parameters, revealing clinically meaningful differences in efficacy based on treatment duration and dosing patterns.

A notable finding is the relationship between treatment duration and clinical outcomes. While the traditional 25-day regimen showed marginal efficacy (RR = 2.29, 95% CI: 1.05-5.00, NNT = 16), extending treatment to 6 weeks yielded improved effects (RR = 3.25, 95% CI: 2.21-4.79, NNT = 7), and 12 weeks showed the highest numerical efficacy (RR = 3.67, 95% CI: 2.60-5.17, NNT = 6). The change in NNT from 16 to 6 represents a clinically important improvement, indicating a substantial reduction in the number of patients needed to treat for one additional successful quit.

However, these differences by treatment duration did not reach statistical significance (χ^2^ = 1.19, df = 2, *p* = 0.55). This lack of statistical significance represents an important limitation and cannot exclude the possibility that observed differences are due to chance. Given the small number of trials in each subgroup (25 days: 4 trials, 6 weeks: 3 trials, 12 weeks: 3 trials), statistical power to detect true differences was likely insufficient. Therefore, while these findings are promising, additional evidence is needed to draw definitive conclusions.

### Sources of Heterogeneity and Impact of Special Populations

The substantial heterogeneity observed in the overall analysis (I^2^ = 88.2%) was primarily driven by the study by Dogar et al.(31) conducted in tuberculosis patients. This study observed an unusually high quit rate in the placebo group (29.7%), markedly different from other studies where placebo quit rates typically ranged from 5-10%. When this study was excluded in sensitivity analysis, the overall effect estimate increased (RR = 3.46, 95% CI: 2.74-4.38) and heterogeneity was completely resolved (I^2^ = 0.0%).

This finding suggests that tuberculosis patients represent a special population with different cessation motivations and health behaviors compared to the general smoking population. The heightened health concerns following tuberculosis diagnosis may have led to high quit rates even in the placebo group. While the Baujat plot did not identify Dogar et al. as contributing substantially to heterogeneity (likely due to its effect estimate being close to null), the leave-one-out analysis clearly demonstrated its unique impact on the pooled results.

### Interpretation of Subgroup Analyses: Balancing Promise and Caution

The complete elimination of heterogeneity observed with extended treatment durations is particularly striking. Both 6-week and 12-week regimens showed perfect homogeneity (I^2^ = 0%), contrasting sharply with substantial heterogeneity in the 25-day regimen (I^2^ = 79%). This pattern suggests that longer treatment durations may provide more consistent treatment effects across different populations and settings.

Similarly, the comparison between fixed-dose regimens (RR = 4.64, 95% CI: 2.60-8.26, I^2^ = 0%, NNT = 5) and declining-dose approaches (RR = 2.45, 95% CI: 1.33-4.50, I^2^ = 88%, NNT = 11) shows substantial differences in both effect size and heterogeneity. While not statistically significant (χ^2^ = 2.22, df = 1, *p* = 0.14), the complete homogeneity in the fixed-dose group suggests high reproducibility of this dosing method.

The meta-regression analysis, while not reaching statistical significance (*p* = 0.217), showed that treatment duration explained 51.3% of between-study heterogeneity. This substantial R^2^ value suggests clinical relevance despite the lack of statistical significance, further supporting the potential importance of treatment duration in optimizing cytisine therapy.

These findings should be interpreted as exploratory and hypothesis-generating. While the observed trends indicate directions for future research, they are insufficient for making definitive clinical recommendations at this time.

### Comparison with Previous Evidence

Our findings build upon and extend the existing cytisine evidence base. Historical meta-analyses have reported varying effect estimates, from RR = 3.98(7) in early analyses to more conservative estimates of RR = 1.30 (Livingstone-Banks et al. 2023) in recent Cochrane reviews. Our intermediate estimate of RR = 2.92 (or RR = 3.46 when excluding the tuberculosis population) likely reflects the inclusion of both traditional and optimized regimens, as well as methodological advances in trial conduct over time.

Importantly, previous meta-analyses have not systematically explored treatment optimization with the granularity presented here. While Puljevic et al. (2024)(10) included only two trials comparing treatment durations, our analysis incorporating seven trials provides more detailed insights into duration-dependent and dose-pattern effects, though definitive conclusions await further evidence.

### Mechanistic Insights and Pharmacological Considerations

The numerically superior performance observed with extended treatment durations aligns with theoretical understanding of nicotine dependence neurobiology. Chronic nicotine exposure leads to nicotinic acetylcholine receptor upregulation and complex neuroadaptive changes that require time to normalize(32). The 25-day regimen may be insufficient for complete receptor resensitization and restoration of normal cholinergic function. Extended treatment may allow for more gradual neurobiological recovery while maintaining pharmacological support during the critical early abstinence period(33).

The complete resolution of heterogeneity with fixed-dose regimens suggests that maintaining steady-state plasma concentrations may be more important than the traditional tapering approach. However, these mechanistic explanations remain speculative given the lack of statistical significance in subgroup comparisons.

### Safety Considerations

The small but statistically significant increase in severe adverse events (RR = 1.12, 95% CI: 1.01-1.24) translates to approximately 12 additional events per 1000 treated patients. Importantly, the safety analysis showed minimal heterogeneity (I^2^ = 5.7%), suggesting a consistent safety profile across different regimens and populations. This favorable safety profile, combined with cytisine’s low cost, supports its use as a first-line treatment option, particularly in resource-limited settings.

### Implications for Clinical Implementation

While the observed numerical trends are compelling, the lack of statistical significance in subgroup comparisons precludes definitive recommendations for changing standard treatment protocols. Clinicians should continue to follow current guidelines while remaining aware of emerging evidence suggesting potential benefits of extended treatment durations. The complete elimination of heterogeneity with 6-week and 12-week regimens provides reassurance about the consistency of treatment effects when using these protocols.

The improvement in NNT from 16 (25-day regimen) to 6 (12-week regimen), if confirmed in future studies, would represent a substantial enhancement in treatment efficiency. Given cytisine’s low cost (approximately $20-30 per course), even extended 12-week regimens would remain highly cost-effective compared to varenicline or combination nicotine replacement therapy.

### Strengths and Limitations

**Strengths** of this systematic review include:

1. Comprehensive search strategy without language restrictions
2. Rigorous methodology following PRISMA guidelines
3. Innovative focus on treatment optimization parameters
4. Detailed exploration of heterogeneity sources through multiple analytical approaches
5. Inclusion of the most recent high-quality trials

**Limitations** must be acknowledged:

1. **Limited statistical power**: Only 7 trials were available for analysis, with even fewer in each subgroup, limiting ability to detect true differences
2. **High overall heterogeneity**: While largely explained by the tuberculosis population study, this suggests unmeasured factors may influence treatment response
3. **Lack of individual patient data**: This prevented exploration of patient-level treatment response modifiers
4. **Limited assessment of publication bias**: With only 7 studies, formal testing for publication bias was not appropriate according to Cochrane guidelines
5. **Insufficient data on long-term outcomes**: Most studies reported outcomes at 24-26 weeks; longer-term effectiveness remains uncertain

### Research Priorities and Future Directions

Our findings highlight critical research priorities:

1. **Adequately powered head-to-head trials**: Direct comparison of 25-day versus 12-week regimens in large, multicenter RCTs is essential to confirm whether observed NNT differences are real
2. **Fixed-dose versus tapering regimens**: Given the striking homogeneity with fixed-dose approaches, comparative effectiveness trials are warranted
3. **Special populations**: Separate trials in populations with comorbidities (excluding them from primary analyses of general smoking populations)
4. **Pharmacokinetic studies**: Investigation of optimal dosing based on cytisine’s half-life and receptor occupancy
5. **Real-world effectiveness**: Implementation studies examining adherence and outcomes with extended regimens in routine practice

### Global Health Implications

For the 1.3 billion tobacco users worldwide, particularly in low- and middle-income countries where cost is a major barrier to cessation treatment, optimizing cytisine therapy represents a crucial public health opportunity. If future trials confirm the superiority of extended regimens, the improvement from NNT = 16 to NNT = 6 would mean helping millions more smokers quit successfully without increasing treatment costs substantially.

### Conclusions

This systematic review and meta-analysis confirms cytisine’s efficacy for smoking cessation and provides exploratory evidence suggesting that treatment optimization—particularly extending duration to 12 weeks and using fixed-dose regimens—may substantially improve outcomes. The complete resolution of heterogeneity when excluding special populations and when analyzing extended treatment regimens provides important insights for future research design.

While the lack of statistical significance in subgroup comparisons precludes definitive clinical recommendations, the magnitude and consistency of observed trends, combined with favorable safety profiles and low treatment costs, strongly support investment in confirmatory trials. Until such evidence emerges, cytisine should be used according to current guidelines, with awareness that ongoing research may soon establish more effective treatment protocols. In the global fight against tobacco-related death and disease, optimizing this affordable and effective treatment remains both a scientific imperative and an ethical obligation.

## Data Availability

All data produced in the present study are available upon reasonable request to the authors

## Acknowledgements

We deeply appreciate to our colleagues in Tokyo Meta Nexus.

## Author contributions

YT: Conceptualization, Methodology, Formal analysis, Writing - Original Draft. KK: Data curation, Investigation. WS: Data curation, Validation. KO: Supervision.

## Data Sharing Statement

The dataset including extracted data from all included studies and R code used for all analyses are available from the corresponding author upon reasonable request. Individual participant data from the original trials were not available to the study authors.

## Figure Legends

**Supplementary Table 1 | Baseline Characteristics of Participants in Included Studies**

Baseline demographic and smoking characteristics of participants enrolled in the seven randomized controlled trials included in the meta-analysis. Data are presented as n (%) for categorical variables and mean ± standard deviation for continuous variables. The Pastorino et al. trial included only current smokers who were also participants in a lung cancer screening program. FTND scores range from 0 to 10, with higher scores indicating greater nicotine dependence. CO levels are measured in parts per million (ppm).

Abbreviations: BMI, body mass index; CO, carbon monoxide; CPD, cigarettes per day; FTND, Fagerström Test for Nicotine Dependence; IQR, interquartile range; NR, not reported; SD, standard deviation.

**Supplementary Table 2 | GRADE Evidence Profile for Cytisine Efficacy**

GRADE (Grading of Recommendations Assessment, Development and Evaluation) evidence profile showing the quality assessment for the primary outcome of biochemically-verified continuous abstinence. The evidence was rated as moderate quality, downgraded one level due to substantial heterogeneity (I^2^ = 92.5%), which was largely explained by treatment duration differences in subgroup analysis. No additional downgrading was applied for study limitations (low risk of bias overall), inconsistency within optimized subgroups, indirectness, imprecision, or publication bias. The moderate quality rating indicates that the true effect is likely to be close to the estimate, but further research may change the confidence in the estimate.

**Supplementary Table 3 | Risk of Bias Assessment Using RoB 2.0 Tool**

Comprehensive risk of bias assessment for all included studies using the revised Cochrane Risk of Bias tool (RoB 2.0). The assessment covers five domains: bias arising from the randomization process, bias due to deviations from intended interventions, bias due to missing outcome data, bias in measurement of the outcome, and bias in selection of the reported result. Overall, 5 studies were rated as low risk of bias, and 2 studies had some concerns, with no studies rated as high risk. The most common concerns related to missing outcome data due to differential dropout rates between treatment arms. This assessment supports the overall reliability of the included evidence and the validity of the meta-analysis results.

**Supplementary Figure 1 | Leave-One-Out Sensitivity Analysis** Leave-one-out analysis examining the effect of excluding each study on the pooled estimate. To avoid double-counting of placebo groups, only the longest follow-up observation from studies reporting multiple timepoints was used. The analysis demonstrates that excluding the Dogar et al. study substantially increases the pooled effect estimate (RR = 3.29, 95% CI: 2.64-4.10) and completely resolves heterogeneity (I² = 0%). Exclusion of any other single study maintains moderate to high heterogeneity (I² ranging from 83% to 91%) with effect estimates remaining relatively stable (RR ranging from 2.12 to 2.54). The red dashed line indicates the reference risk ratio of 1.0. This analysis identifies the Dogar et al. study, conducted in tuberculosis patients, as the primary source of heterogeneity in the overall meta-analysis.

**Supplementary Figure 2 | Baujat Plot for Heterogeneity Diagnostics** Baujat plot visualizing each study’s contribution to overall heterogeneity (x-axis) and influence on the pooled effect estimate (y-axis). Studies positioned further to the right contribute more to heterogeneity, while those higher on the y-axis have greater influence on the overall result. To avoid double-counting of placebo groups, only the longest follow-up observation from studies reporting multiple timepoints was used. The plot reveals that while Dogar et al. has modest influence on the overall result (due to its effect estimate being close to null), it does not appear as a major contributor to heterogeneity in this visualization. This apparent discrepancy with the leave-one-out analysis reflects the mathematical properties of the Baujat plot, which considers both effect size deviation and precision. Studies are labeled twice in the plot to enhance readability of overlapping points.

**Supplementary Figure 3 | Meta-regression of Treatment Duration and Effect Size** Meta-regression analysis exploring the relationship between cytisine treatment duration (in weeks) and treatment effect (log risk ratio). Each circle represents a study, with size proportional to study weight. The blue line shows the fitted regression model with 95% confidence interval (shaded area). Studies using 25-day regimens were coded as 3.57 weeks for analysis. The positive slope (β = 0.069, 95% CI: −0.041 to 0.179) suggests a trend toward increased efficacy with longer treatment duration, though this did not reach statistical significance (p = 0.217). Treatment duration explained 51.3% of between-study heterogeneity (R² = 51.3%), suggesting clinical relevance despite lack of statistical significance. The Dogar et al. study appears as an outlier below the regression line, consistent with its unique patient population. To maximize information about the duration-response relationship, all data points from studies reporting multiple timepoints (ORCA-2, ORCA-3, and Pastorino) are included.

