## Supplementary figures and images for "Cytisine for smoking cessation: A systematic review and meta-analysis suggesting potential benefits of extended treatment protocols"

### Supplementary figures

#### Slide 1
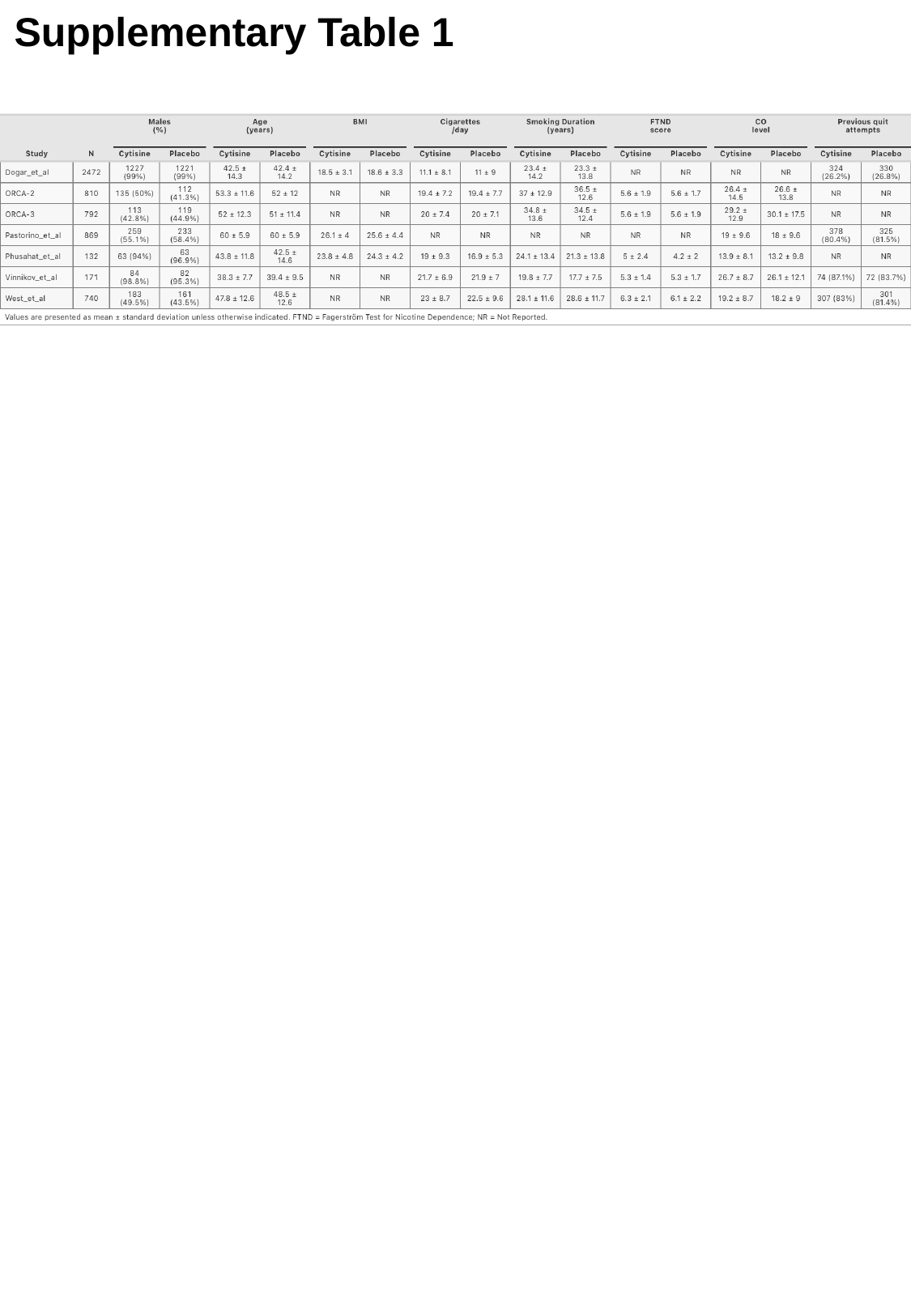

Supplementary Table 1

#### Slide 2
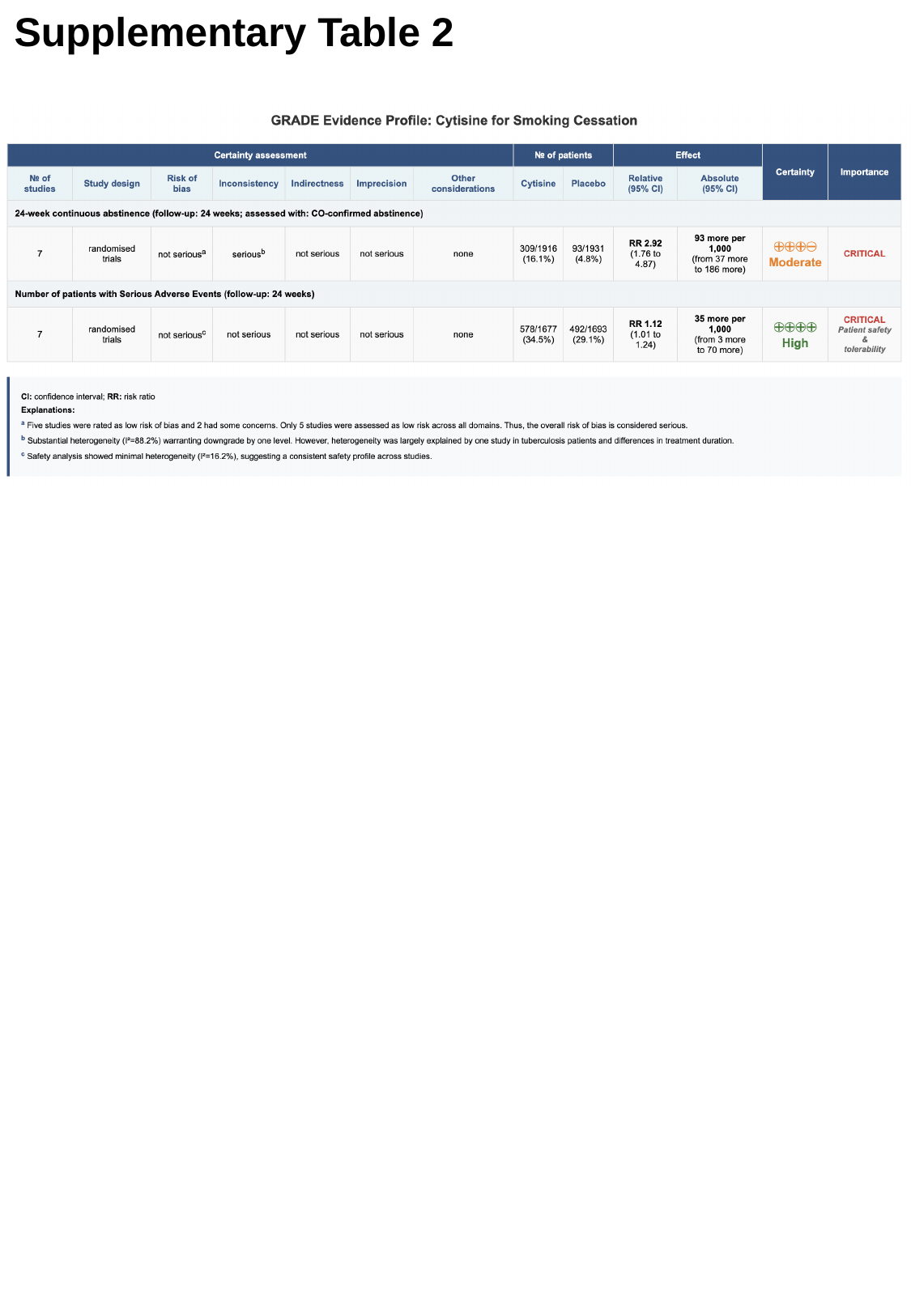

Supplementary Table 2

#### Slide 3
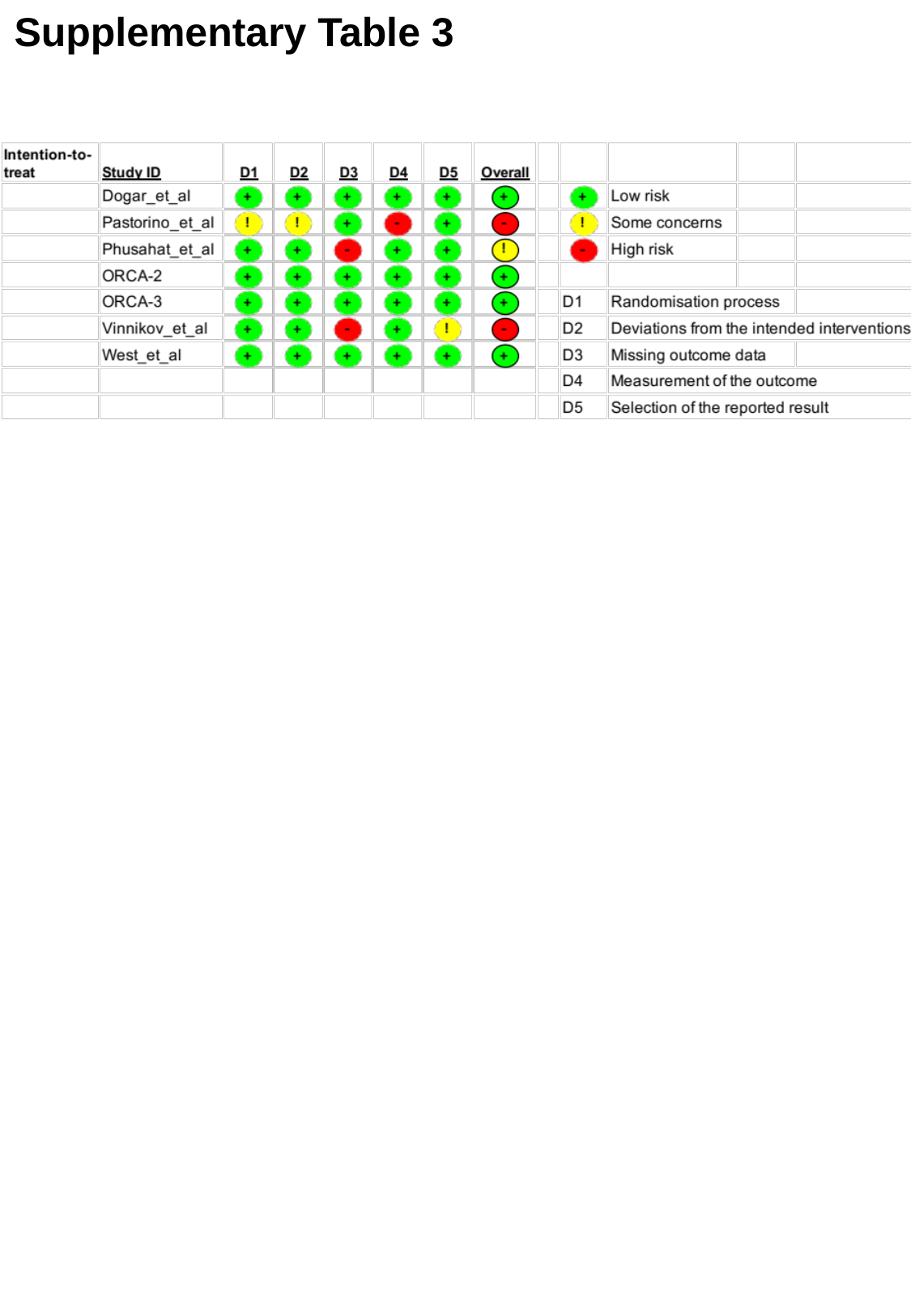

Supplementary Table 3

### Supplementary tables

#### Slide 1
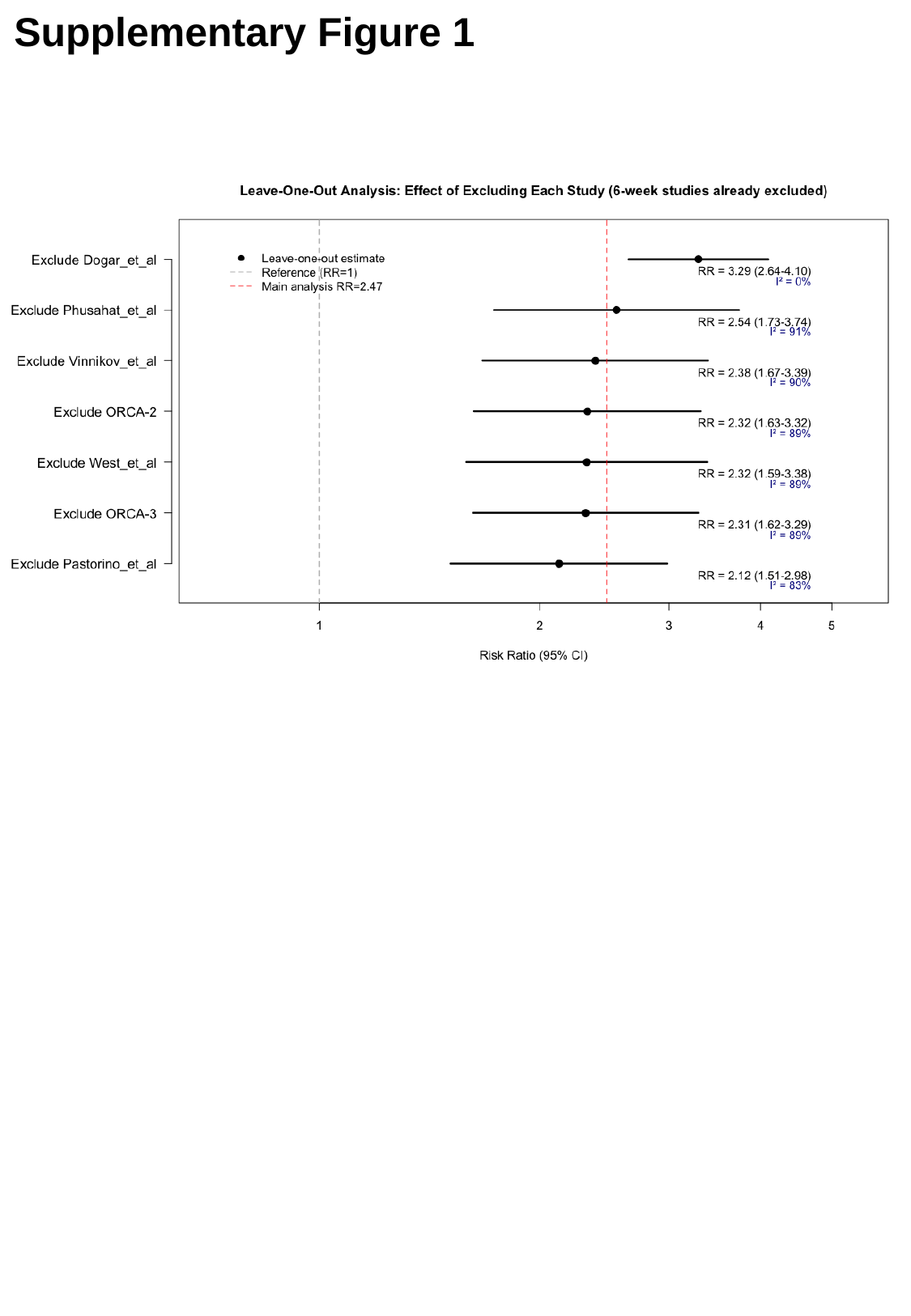

Supplementary Figure 1

#### Slide 2
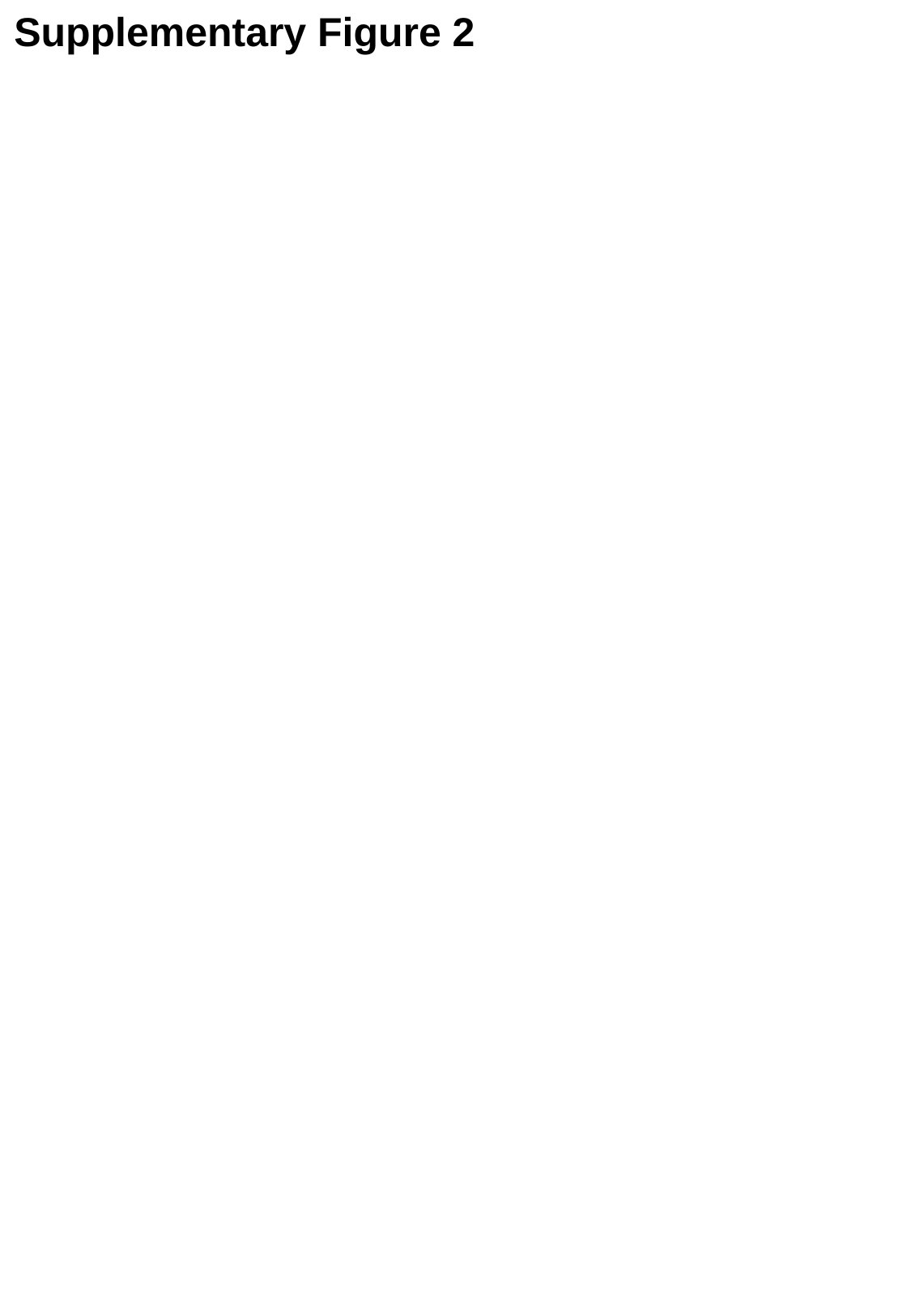

Supplementary Figure 2

#### Slide 3
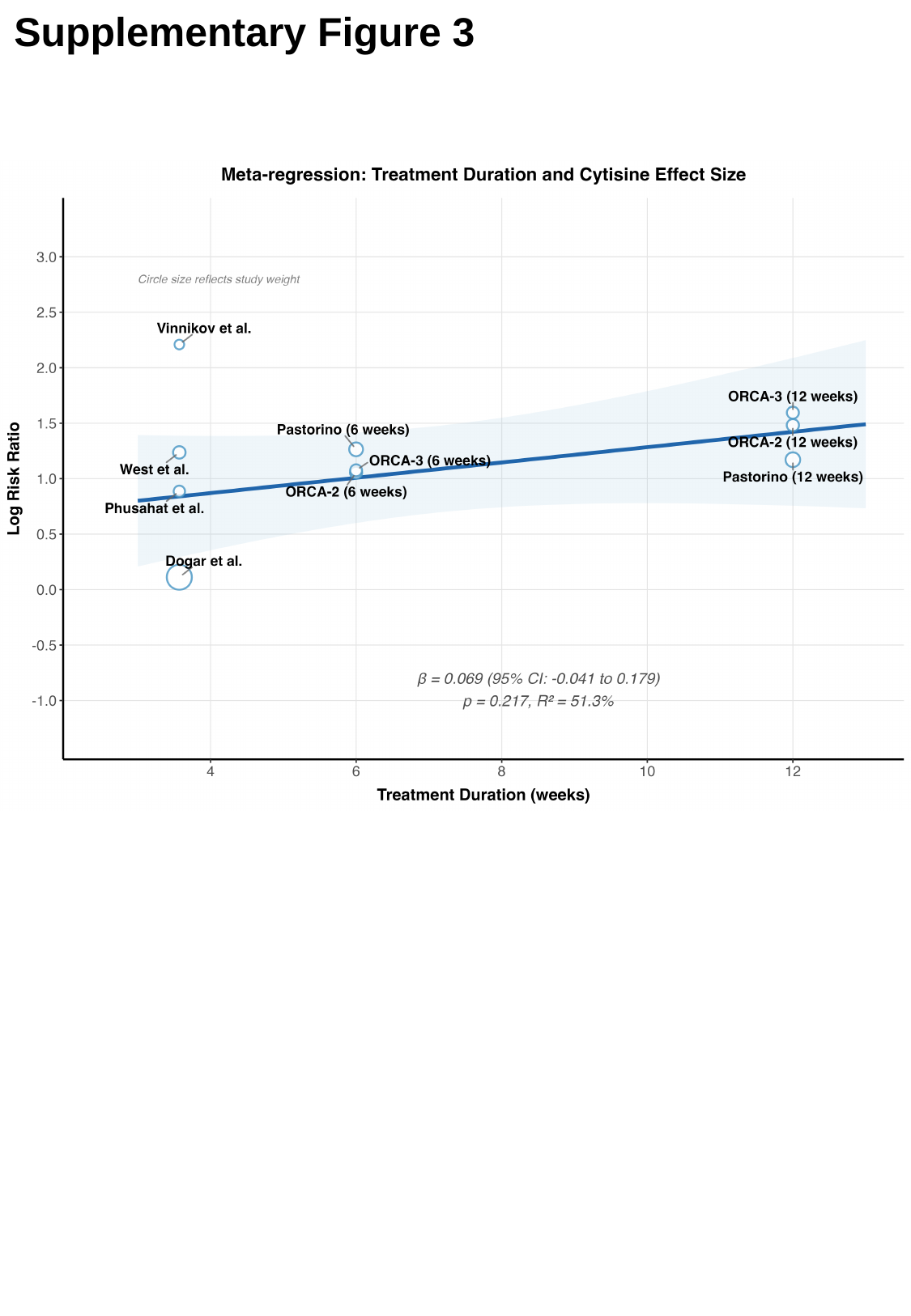

Supplementary Figure 3
